# AS04 in a bivalent HPV vaccine drives superior cross-protective antibody response by increased NOTCH signaling of cDC1 leading to increased proliferation of adaptive immune cells

**DOI:** 10.1101/2025.02.25.25322835

**Authors:** Valentino D’Onofrio, Ana-Carolina Santana, Marthe Pauwels, Gwenn Waerlop, Anthony Willems, Fien De Boever, Peter Sehr, Tim Waterboer, Isabel Leroux-Roels, Ashish Sharma, Rafick Pierre Sékaly, Geert Leroux-Roels

## Abstract

**Introduction:** Cervarix® and Gardasil® are two HPV vaccines with differing antigen and adjuvant compositions. Gardasil-4 contains HPV types 6, 11, 16 and 18 type-specific L1 viral like particles (VLPs) formulated with amorphous AlHO9PS-3 adjuvant, while Cervarix targets HPV types 16 and 18 using AS04 (Al(OH)3 + TLR4 agonist MPL) to enhance immune response and cross-protection against other high-risk HPV types, not included in the vaccine.

**Methods:** To investigate mechanisms of cross-neutralizing potential of Cervarix, six monozygotic twins (12 females aged 9-13 years) were vaccinated with either Cervarix or Gardasil-4 (2 doses, 6 months apart). Serum neutralizing antibody titers against HPV 6,16,18,31,33,45,52, and 58, were assessed pre-vaccination and 7 days post-second dose. Multi-omic single cell RNA and ATAC sequencing of PBMCs were performed at the latter timepoint.

**Results:** Cervarix generated higher cross-neutralizing antibody titers than Gardasil-4. Higher frequencies of plasmacytoid (pDC) and conventional dendritic cells (cDC1, cDC2), CD4+ T effector memory (Tem) and B memory cells were also observed after Cervarix. Cervarix-vaccinated subjects showed increased DC-to-CD4+ Tem and B memory cell signaling, through increased antigen presentation and upregulation of NOTCH pathway. Gene Set Enrichment Analysis indicated enhanced pathways related to cell migration and NOTCH2 signaling in DCs and cell cycling/RNA translation in CD4+ T and B cells, correlating positively with cross-neutralizing antibody titers. Increased chromatin accessability in genes related to NOTCH signaling in cDC1 was also observed. Engagement of MHC and NOTCH induced FOS in CD4+ Tem cells and BCL2 in B memory cells, supporting proliferative and anti-apoptotic states. This also resulted in an increase in Th2 cells in Cervarix-vaccinated subjects, and increased IgG4 expression in B memory cells.

**Conclusion:** Increased DC signaling, including NOTCH, through AS04 in Cervarix supports cell survival and sustained RNA translation in adaptive immune cells, 7 days post-vaccination, especially memory T and B cells. This increased cell metabolism and activation may enhance cell maturation of adaptive immune cells, providing a mechanism triggered by Cervarix that can lead to improved cross-reactivity.

## Introduction

Human papillomavirus (HPV) is one of the most prevalent sexually transmitted infections worldwide, causing cervical cancer and contributing to other malignancies, including vulvar, vaginal, anal, penile, and oropharyngeal cancers (1). Notably, over 99% of cervical cancer cases can be attributed to persistent infection with high-risk HPV types, particularly HPV16 and HPV18, which together account for approximately 70% of global cases (2). Additionally, non-oncogenic HPV types, such as HPV6 and HPV11, are associated with benign anogenital and non-genital warts, underscoring HPV infections as a significant public health concern (1, 3).

To address this significant health burden posed by HPV-related diseases, prophylactic HPV vaccines have been developed to prevent infection and associated malignancies (4). Currently, three globally licensed HPV vaccines are based on the self-assembly properties of the HPV-type specific major capsid protein L1, into virus-like particles (VLPs) (4). While all licensed vaccines are L1- containing VLPs, they differ in their antigen content and adjuvant formulations. Cervarix® (GlaxoSmithKline Vaccines) is a bivalent vaccine targeting HPV16 and HPV18, and is formulated with the Adjuvant System 04 (AS04), comprising aluminum hydroxide salts (Al(OH)_3_) and the Toll- like receptor 4 (TLR4) agonist 3-O-desacyl-4’-monophosphoryl lipid A (MPL) (5). Gardasil-4® (Merck) is a quadrivalent vaccine formulated with amorphous aluminum hydroxyphosphate sulfate (AlHO9PS-3) as its adjuvant, targeting HPV16, HPV18, and the low-risk types HPV6 and HPV11 (6). Gardasil-9® (Merck) maintains the same aluminum-based adjuvant but extends Gardasil-4’s protection by including five additional high-risk HPV types (31, 33, 45, 52, and 58), which collectively account for approximately 20% of cervical cancers (7).

While HPV vaccines are highly effective and safe in preventing persistent HPV-infections and precancerous cervical lesions caused by high-risk HPV types (8, 9), Cervarix and Gardasil-4 have demonstrated differences in their immunological profiles (10–13). Both vaccines elicit high levels of neutralizing serum antibodies against vaccine-specific HPV types (14). However, Cervarix induces higher HPV16 and HPV18 antibody titers, greater frequencies of HPV16/18-specific memory B cells, and more robust antibody-dependent complement activation compared to Gardasil-4 (10, 15–17). Both vaccines confer a differential degree of cross-protection by inducing cross-protective neutralizing antibodies towards phylogenetically related, non-vaccine HPV types within the Alpha- papillomavirus species group A9 (HPV16-like: 31, 33, 35, 52, 58) and A7 (HPV18-like: 39, 45, 59, 68) (18). Cervarix elicits broader cross-neutralization capabilities and higher magnitudes of cross- protective antibodies towards HPV31, 33, 45, and 52, compared to Gardasil-4, with differences in magnitude decreasing with increasing age (10, 13, 15, 17, 18). The superior cross-protective efficacy of Cervarix can be attributed to its AS04 adjuvant. In preclinical models, AS04 directly stimulates dendritic cells, thereby enhancing antigen presentation and promoting T-cell activation (19, 20). However, the molecular mechanisms underlying the broader cross-neutralization capacities of Cervarix remain to be further elucidated.

This study aimed to investigate the molecular mechanisms by which AS04 induces cross-protective antibodies against closely related HPV types not included in Cervarix, in comparison to Gardasil-4. In PBMCs collected seven days after the second vaccine dose, we confirmed an increased breadth of neutralizing antibodies. Additionally, we observed enhanced conventional Dendritic Cells type 1 (cDC1) signaling, including upregulated NOTCH signaling, as evidenced by increased gene expression and chromatin accessibility. This activation was associated with active cell proliferation seven days after the second dose, contributing to the improved maturation of adaptive immune cells.

## Results

### AS04 enhances neutralizing antibody responses

To investigate the effect of AS04 on antibody responses and its molecular mechanisms, six female homozygotic twins (n = 12), were vaccinated with either Cervarix, containing AS04, or Gardasil-4, containing alum. All participants were naive for HPV and received the vaccine at the recommended age of 9-13 years in Belgium. Each regimen consisted of 2 doses, 6 months apart (day 0 and day 180) (**Figure 1A**). Serum was collected at day 0 and 7 days post-second dose (day 187), while blood for peripheral blood mononuclear cell (PBMC) isolation was collected at day 187 only. The neutralizing antibody response was measured using a pseudovirion-based neutralization assay (21). Both vaccines target the L1 protein of HPV16 and 18 and successfully induced neutralizing antibodies (**Figure 1B**), with a marked increase observed from day 0 to day 187 (**Supplementary Table 1**). Gardasil-4 additionally targets HPV6 and 11 (type 11 not measured). Neutralizing antibodies against HPV31, 33, 52 and 58 (closely related to HPV16) and HPV45 (closely related to HPV 18) were also increased by day 187 (**Figure 1B**). On day 187, neutralizing antibody titers against HPV18 and 45 were significantly higher in participants vaccinated with Cervarix, consistent with previous studies demonstrating Cervarix‘s superior response to HPV18 compared to Gardasil-4 (**Figure 1C**, **Supplementary Table 1**). Conversely, titers against HPV6 were significantly higher following Gardasil-4 vaccination, as this antigen is included in Gardasil-4 but not in Cervarix (**Figure 1C**). While titers against HPV16 and other related types (31, 33, 52 and 58) showed no significant differences between vaccines, a trend favoring Cervarix was observed (**Figure 1C**). Overall, these findings confirm that Cervarix has a greater capacity to enhance cross-protective antibody responses compared to Gardasil-4, thereby broadening the immune response. On day 187, no differences in serum cytokine concentrations were observed between vaccines (**Figure 1E, Supplementary Table 1**).

**Figure 1.**
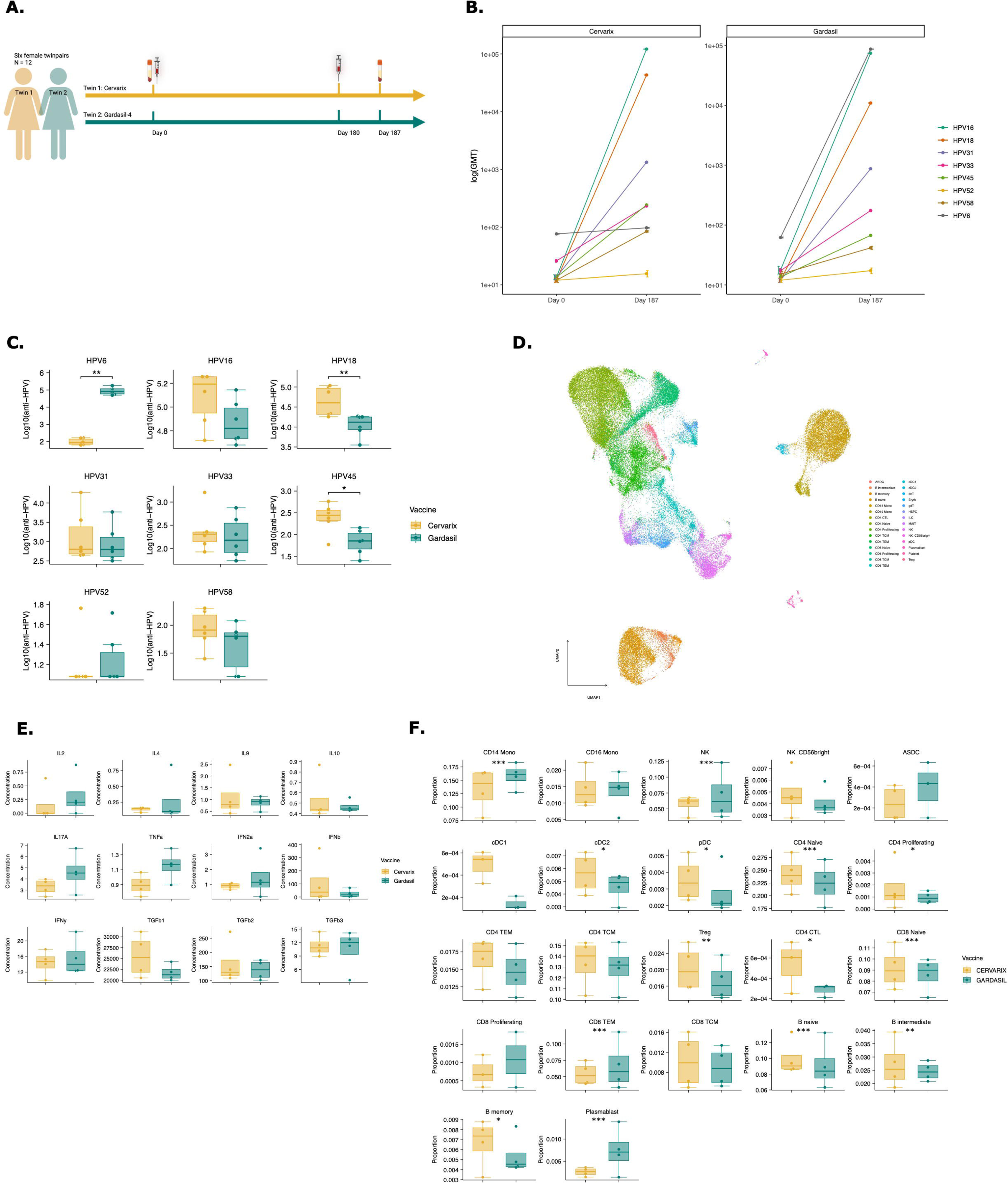
Cervarix induces higher neutralizing antibody titers against closely related HPV types and enhances DC and Memory cell frequencies in peripheral blood. **A.** Study design: Six female homozygotic twins (n = 12) were vaccinated with either Cervarix, containing AS04, or Gardasil-4, containing alum. All participants were HPV-naive and received the vaccine at the recommended age of 9-13 years in Belgium. Each regimen consisted of two doses administered six months apart (day 0 and day 180). Serum samples were collected on day 0 (pre-vaccination) and seven days after the second dose (day 187), while blood for PBMC isolation was collected on day 187 only. **B.** Neutralizing antibody titers: neutralizing antibody titers against HPV6, 16, 18, 31, 33, 45, 52 and 58 were measured using a pseudovirion-based neutralizing assay in all participants at both timepoints. Geometric mean titers (GMT) were calculated for each HPV type and vaccine. The line graph presents log-transformed GMT on day 0 and day 187 for each vaccine separately, with colored line representing specific HPV types. **C.** Neutralizing antibody levels on day 187: Boxplots display the median (IQR) log-transformed neutralizing antibody titers on day 187 for each HPV type, grouped by vaccine (orange: Cervarix, green: Gardasil-4). **D.** Cytokine concentrations: Boxplots show the median (IQR) of log-transformed cytokine concentrations measured in serum from day 187 grouped by vaccine. E. Single cell RNA sequencing analysis: A total of 79,817 cells were analyzed using single-cell RNA sequencing. The UMAP plot visualizes all single cells after dimensionality reduction, clustering, and annotation. F. Cell frequencies: Boxplots show the median (IQR) of relative cell frequencies per vaccine. Abbreviations: CD14 Mono: classical CD14+ monocytes; CD16 Mono: non-classical CD16+ monocytes; NK: Natural Killer cells; ASDC: AXL+ Siglec-6+ dendritic cells; cDC1/cDC2: conventional dendritic cells type 1 or type 2; pDC: plasmacytoid dendritic cell; CD4 TEM: CD4+ T effector memory cells; CD4 TCM: CD4+ T central memory cells; Treg: regulatory T cells. CD4 CTL: CD4+ cytotoxic T cell; CD8 TEM: CD8+ T effector memory cells; CD8 TCM: CD8+ T central memory cells; ILC: innate lymphoid cells; GMT: geometric mean titer; HPV: human papilloma virus; ns: not significant. * p-value <0.05, ** p- value <0.01, *** p-value <0.001 by Wilcoxon rank test.

### AS04 enhances dendritic cell recruitment

Next, we investigated the differential effect of AS04 on transcriptional and epigenetic changes in PBMCs. Four twins (n = 8) representative of the differences in neutralizing antibody titers across all types of HPV, were selected by calculating the Euclidian distance (**Supplementary Figure 1A**). Nuclei from these eight PBMC samples collected on day 187 were subjected to multi-ome single cell RNA and ATAC sequencing (10X Genomics). In total, 79,817 cells were sequenced, and 71,767 high-quality cells were retained for analysis, including clustering and annotation using the Azimuth PBMC reference (**Figure 1D, Supplementary** Figure 1). Statistically significant differences in relative frequencies of innate and adaptive immune subsets were found for most cells using Wilcoxon Rank-sum test (**Figure 1F**). There was a lower proportion of classical CD14+ monocytes (odds ratio (OR): 0.81; [0.77 – 0.84] 95%CI) and cytotoxic Natural Killer (NK) cells (OR: 0.77; [0.72 – 0.82] 95%CI) after Cervarix compared to Gardasil-4 (**Supplementary Table 2**). Cervarix showed a higher proportion of conventional dendritic cells type 2 (cDC2) (OR: 1.25; [1.01 – 1.56] 95%CI) and plasmacytoid dendritic cells (pDC) (OR: 1.18; [0.90 – 1.55] 95%CI); these innate immune subsets are known for enhanced antigen presentation. Although not statistically significant, Cervarix also induced higher frequencies of conventional DCs type 1 (cDC1) (**Supplementary Table 2**). A higher proportion of memory B cells was observed following Cervarix vaccination (OR: 1.21; [1.00 – 1.47] 95%CI), but a decreased proportion of plasmablasts (OR: 0.33; [0.26 – 0.42] 95%CI)compared to Gardasil-4 (**Figure 1F, Supplementary Table 2**). There was also a trend towards a higher proportion of effector and central memory CD4+ T cells after Cervarix vaccination (**Figure 1F**, **Supplementary Table 2**).

### AS04 enhances signaling from DCs to adaptive immune cells and drives cell survival

Since we observed increased frequencies of DC subsets in both vaccine regimen, we next examined the molecular signals mediating communication from DCs to adaptive immune cells. Using the CellChat package, we analyzed cell-cell interactions, defined as ligand-receptor interactions inferred from the gene expression matrix, between innate and adaptive immune cells after Cervarix and Gardasil-4 vaccination. A comparison of interaction counts between innate and adaptive immune cells revealed more interactions following Cervarix vaccination (2160 versus 2096 predicted ligand-receptor interactions after Cervarix and Gardasil-4, respectively) (**Supplementary Figure 2A**). Specifically, interactions between cDC1/pDC and adaptive immune cells were more frequent after Cervarix, whereas interactions between AXL+ SIGLE6+ DCs (ASDCs) and adaptive immune cells predominated after Gardasil-4 (**Figure 2A**).

**Figure 2.**
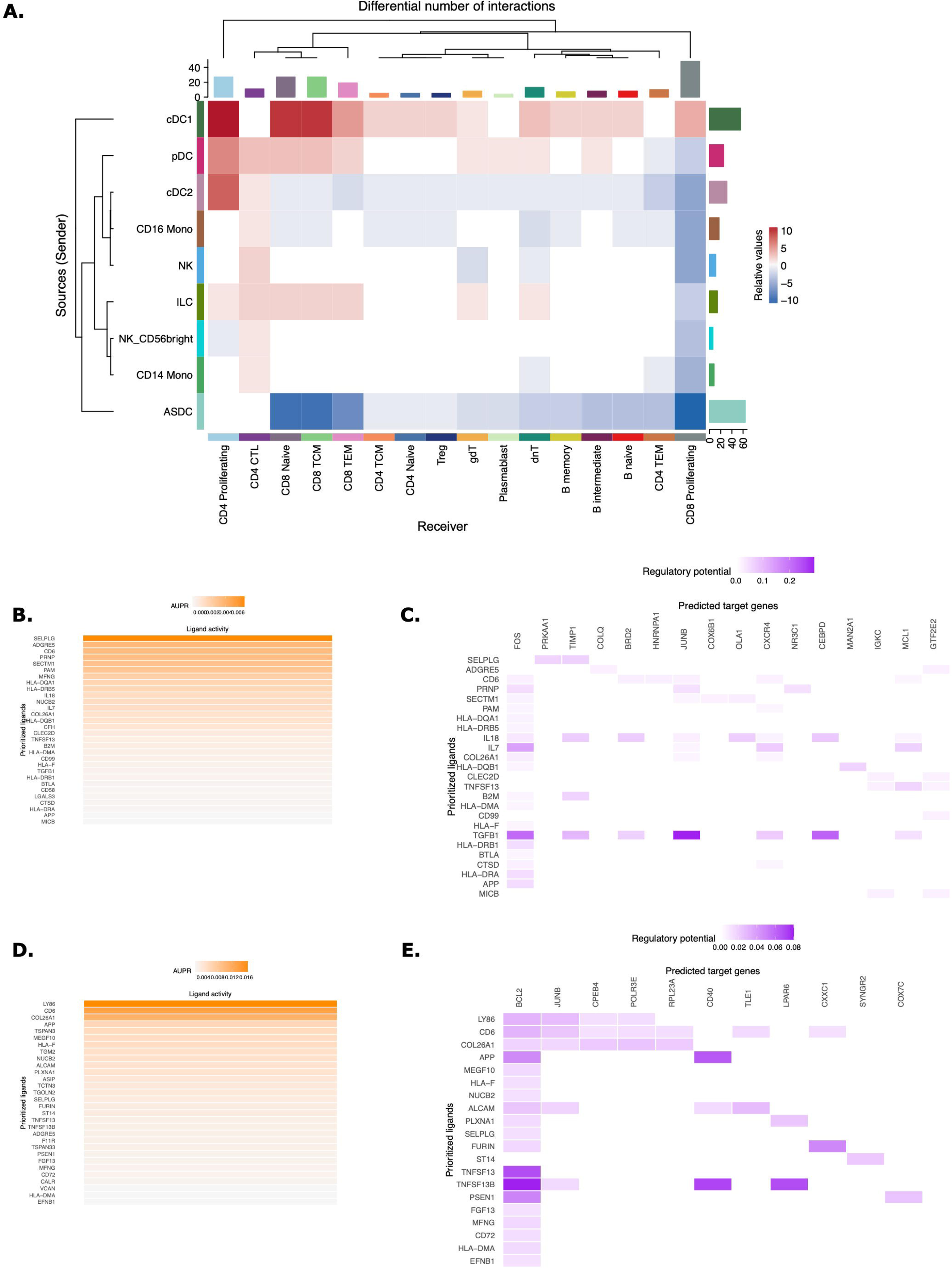
Enhanced signaling by DCs via NOTCH after Cervarix vaccination. **A.** Heatmap comparing the differential number of interactions between Cervarix and Gardasil-4. Rows represent sending cells (y-axis), while columns indicate receiving cells (x-axis). Red denotes more signaling in Cervarix, while blue indicates increased signaling in Gardasil-4. The heatmap is clustered based on the differential number of interactions. Bar plots along the rows and columns indicate the total number of signals sent or received, respectively. **B-E.** NicheNet analysis of DC signaling to CD4+ T cells and B memory cells. **B.** Ligand activity plot highlighting the most important ligands sent by cDC1 and pDC to CD4+ TEM cells. **C.** Target gene heatmap showing the expression of genes in CD4+ TEM cells (columns) influenced by the top ligands (rows). **D.** Ligand activity plot highlighting the most important ligands sent by cDC1 and pDC to memory B cells. **E.** Target gene heatmap showing the expression of genes in memory B cells (columns) influenced by the top ligands (rows). Abbreviations: CD14 Mono: classical CD14+ monocytes; CD16 Mono: non-classical CD16+ monocytes; NK: Natural Killer cells; ASDC: AXL+ Siglec-6+ dendritic cells; cDC1/cDC2: conventional dendritic cells type 1 or type 2; pDC: plasmacytoid dendritic cell; CD4 TEM: CD4+ T effector memory cells; CD4 TCM: CD4+ T central memory cells; Treg: regulatory T cells. CD4 CTL: CD4+ cytotoxic T cell; CD8 TEM: CD8+ T effector memory cells; CD8 TCM: CD8+ T central memory cells; ILC: innate lymphoid cells; gdT: gamma delta T cells; dnT: double negative T cells; AUPR: area under the precision recall curve.

For both vaccines, pDCs were identified as the primary ligand-sending cells, with MHC-I and MHC- II being the most frequently used ligands (**Supplementary Figure 2B-C**). However, after Cervarix vaccination, cDC1 and cDC2 played a more prominent role in sending signals. These signals included ligands that are important in recruitment, adhesion, and activation of T cells (APP, NEGR, SELPLG, SEMA3, CD45, CD86, PECAM1, ADGRG, CD99, CLEC, BAFF). Interestingly, several of these ligands, BAFF, CD86, PECAM1, impact B and T cell survival. While ASDCs were more involved after Gardasil-4, similar ligands were sent. Additionally, MHC-II presented by cDC1 and cDC2 was more important after Cervarix vaccination compared to Gardasil-4 (**Supplementary Figure 2C-D**). Unsurprisingly, CD8+ T cells were the primary recipients of these signals for both vaccines (**Supplementary Figure 2D-E**).

To further investigate the functional outcomes of these interactions, we used NicheNet to identify genes differentially expressed after Cervarix vaccination in recipient cells (CD4+ T effector memory (CD4+ TEM) and Memory B cells) upon receiving these signals. Similarly signaling from DCs to CD4+ TEM cells and memory B cells was evidenced by the activity of several ligands involved in recruitment and adhesion (SELPLG, ADGRE5, APP) of adaptive immune cells and allowing for stable synapses needed for effective communication.

Again, ligands, including MFNG (NOTCH signaling), CD6 (co-stimulation), IL7 (survival), and BAFF (B cell activating factor), that stimulate cell proliferation and survival signals in CD4+ TEM cells were found, (**Figure 2B**). These signals promoted the expression of several genes critical for CD4+ TEM cell survival, such as FOS, JUNB, MCL1, NR3C1, and TIMP1 (**Figure 2C**). Furthermore, signaling from DCs to memory B cells included cell proliferation and survival ligands such as MFNG (NOTCH signaling), BAFF, and PSEN1 (NOTCH signaling) (**Figure 2D**). These ligands further enhanced memory B cell survival by inducing the expression of survival-associated genes like BCL2, CD40 and JUNB (**Figure 2E**). Overall, these findings demonstrate that, in addition to increased antigen presentation by DCs, these cells signal the recruitment of CD4+ TEM cells and B memory cells, driving their proliferation and survival, and promoting enhanced adaptive immune response.

### AS04 enhances NOTCH signaling through gene expression in cDC1, stimulating cell cycling in memory CD4+ and B cells, which correlates with breadth of neutralizing antibodies

To further investigate the function of innate immune cells, differentially expressed genes (DEG) in each cell type were identified and analyzed using gene set enrichment analysis (GSEA) (**Supplementary Table 3**). Notably, enrichment of the Slit-Robo signaling pathway, and increased expression of genes like TRIO and ARGHAP24, which are involved in axon guidance and cell migration, were observed across all innate cell types following Cervarix vaccination, suggesting enhanced recruitment of these cells (**Supplementary Figure 3C**). Additionally, ribosomal proteins were among the most differentially expressed genes, and genes related to RNA translation, cell metabolism and cell proliferation pathways were enriched in the Cervarix group, which indicates an enhanced cellular machinery for protein synthesis and can support a more robust immune response by facilitating the activation and proliferation of immune cells (**Supplementary Figure 3D, Supplementary Table 3**). In cDC1 and pDCs, this was accompanied by increased expression of transcriptional genes involved in RNA translation, protein folding and cell-cycling, -development, and -apoptosis. Notably, TAO Kinase 1 (TAOK1) and Catenin Beta 1 (CTNNB) were upregulated in cDC1 (**Figure 3A, Supplementary Figure 3A**), while Ubiquitously Expressed Prefoldin-Like Chaperone (UXT) and Eukaryotic Translation Initiation Factor 3 (EIF3) were upregulated in pDCs (**Figure 3B**). Consistent with previous findings, genes and transcription factors involved in NOTCH2 signaling, including NOTCH2, MAML2, and RBPJ (**Figure 3C, Supplementary Figure 3B**) showed upregulated average gene expression after Cervarix. Moreover, NOTCH2 signaling, an important regulator of Th2 immunity, was one of the three enriched pathways identified in cDC1 (**Figure 3F**). These findings confirm an increased frequency of innate immune cells, particularly cDC1s (0.03% vs. 0.01%), along with their increased activation and signaling by NOTCH2.

**Figure 3.**
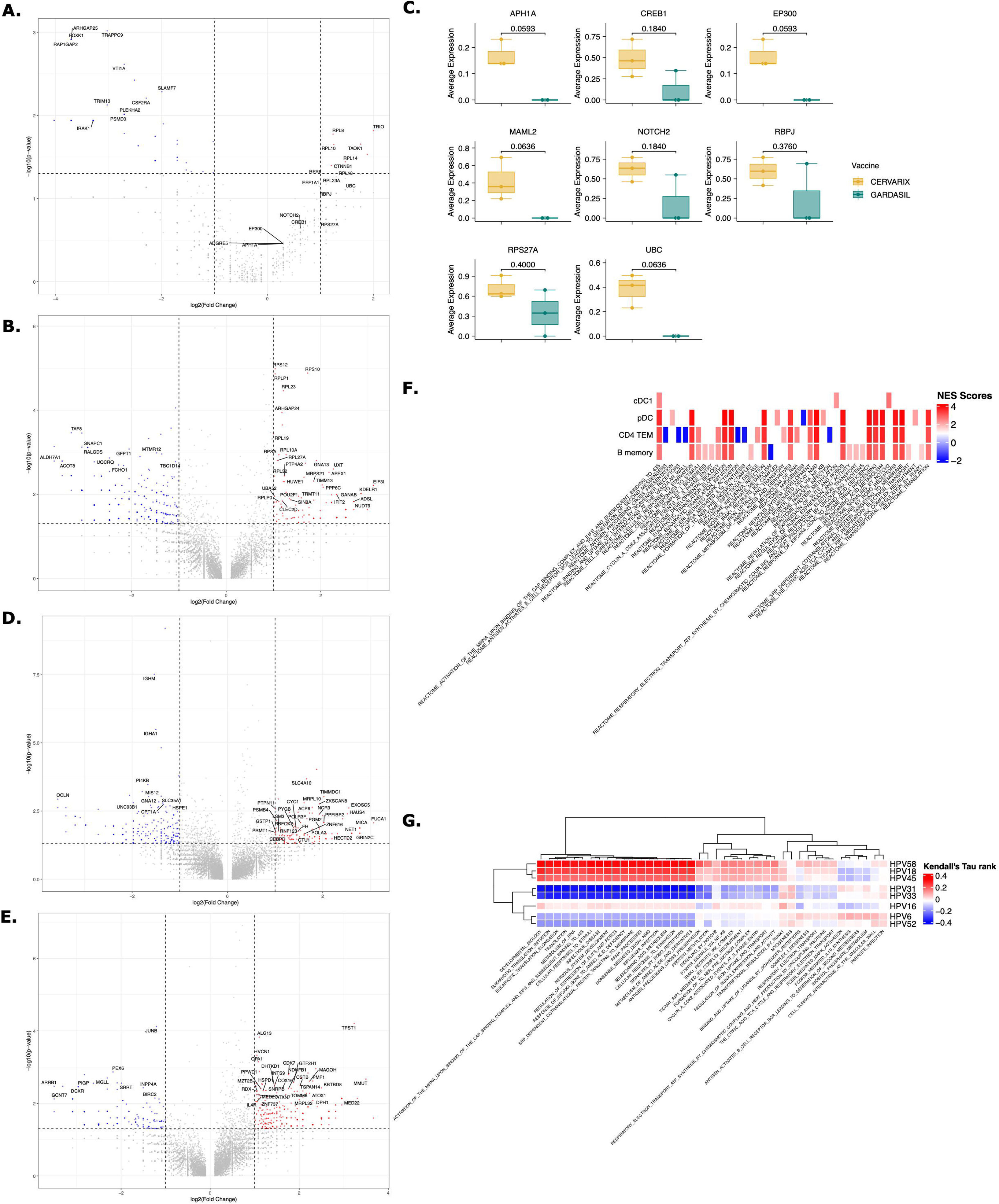
Cervarix induces transcriptional responses in DCs and memory CD4+ T and B cells that correlate with antibody titers. A-B. Volcano plots of differentially expressed genes (DEGs) for cDC1 (**A**), pDC (**B**). **C.** Boxplot showing the median (IQR) of average gene expression of NOTCH2 signaling pathway genes in cDC1, grouped by vaccine. **D-E.** Volcano plots of differentially expressed genes (DEGs) for CD4+ T cells (**D**) and B cells (**E**). F. Heatmap of normalized enriched scores (NES) for significantly enriched reactome pathways in cDC1, pDC, CD4+ TEM, and memory B cells. **G.** Heatmap showing the correlation between significantly enriched pathways and HPV- type-specific neutralizing antibody titers, with color representing the Kendall’s Tau rank. Abbreviations: cDC1: conventional dendritic cells type 1; pDC: plasmacytoid dendritic cell; CD4 TEM: CD4+ T effector memory cells; NES: normalized enrichment score. * p-value <0.05, ** p- value <0.01, *** p-value <0.001 by Wilcoxon rank test.

Ribosomal genes were also the most overexpressed genes in both CD4+ T cells and B cells following Cervarix, compared to Gardasil-4 (**Figure 3D-E, Supplementary Table 3**). Cervarix increased the expression of genes involved in RNA translation, cell metabolism and cell proliferation pathways in all CD4+ T cells and B cells (**Supplementary Figure 3E-F, Supplementary Table 3**). Interestingly, and similar to the cell-cell communication findings, transcription factors and genes involved in cell cycling, cell survival and anti-apoptosis such as CDK7, CYC1, MICA, and OPA1, were among the top expressed genes in CD4+ TEM and memory B cells (**Figure 3D-E**). Indeed, mRNA translation, protein folding, and cell cycling were pathways significantly enriched after Cervarix across cell types. Moreover, pDCs and cDC1 showed enrichment of cell recruitment and cell signaling pathways, while CD4+ TEM and memory B cells showed enrichment of cell survival and proliferation pathways (**Figure 3F**).

We next sought to determine whether the identified pathways in cDC1, pDC, CD4+ TEM cells and B memory cells correlated with higher neutralizing antibody titers, independent of the vaccine. First, Module scores, which is the average expression level of all genes of the respective pathway, for all significantly enriched pathways were calculated. Then, Kendall’s rank correlation coefficients were calculated between the module score and neutralizing antibody titers for various HPV types (**Supplementary Table 4**). Hierarchical clustering based on pairwise distances was performed, and the resulting dendrogram was used to order rows in the heatmap (**Figure 3G**). Clustering revealed the two groups of HPV types: HPV18-45-58, and HPV6-16-31-33-52 based on correlation coefficients, which mostly correspond to the A7 (HPV18-45) and A9 (HPV16-31-33-52-58) HPV groups. The pathways upregulated after Cervarix vaccination, primarily those related to cell cycle and RNA translation of CD4+ TEM and memory B cells, positively correlated with neutralizing antibody titers for HPV18-45-58, aligning with the higher neutralizing antibody titers observed for these types after Cervarix. Ribosomal gene expression in all cells and Slit-Robo signaling in pDCs, CD4+ TEM and memory B cells, also showed weaker positive correlations with neutralizing antibody titers for other HPV types. Conversely, pathways primarily associated with immune responses, such as NFκB signaling and B cell receptor activation, were upregulated in CD4+ TEM after Gardasil-4 vaccination and positively correlated with neutralizing antibody titers for HPV6-16-31-33-52. Interestingly, NOTCH2 signaling, correlated positively with HPV18-45-58 and HPV16.

To further explore differences in gene expression based on the breadth of neutralizing antibody responses, subjects were categorized as “high breadth responders” (n=4 (Cervarix: n=3, Gardasil- 4: n =4)) and “low breath responders” (n=4 (Cervarix: n=1, Gardasil-4: n =3)) based on the sum of neutralizing antibody titers across all HPV types, with the median serving as threshold. Differential gene expression analysis (**Supplementary Figure 4A-C, Supplementary Table 4**) and GSEA identified similar enriched pathways (**Supplementary Table 4**). High breadth responders showed increased ribosomal gene expression and enrichment of pathways related to RNA translation, cell metabolism and development in DCs (**Supplementary Figure 4D**), CD4+ T cells (**Supplementary Figure 4E**), and B cells (**Supplementary Figure 4F**).

These findings suggest that Cervarix promotes cell differentiation and maturation, by DC signaling, until at least day 7, with an increased reliance on memory cells compared to Gardasil-4. Moreover, NOTCH2 signaling by cDC1, leads to enhanced cell proliferation and development in CD4+ TEM cells and memory B cells. The increased signaling and cell proliferation contribute to a broader and more cross-neutralizing humoral antibody response.

### AS04 increases chromatin accessibility of NOTCH-related genes in cDC1s

To investigate epigenetic changes in DCs induced by AS04, we performed single cell ATAC sequencing on PBMCs collected on day 7 after the second dose. In cDC1 and pDCs, we analyzed genomic regions with significantly increased chromatin accessibility, which may indicate prior training of these cells. The top 200 differentially accessible regions (DARs) in Cervarix compared to Gardasil-4 are shown in **Figure 4A** (**Supplementary Table 5**). Interestingly, while most differences in NOTCH-related gene regions were subtle (**Figure 4B**), one cluster of genes exhibited increased accessibility after Cervarix. This cluster included promotor regions of EGR4, HES5, HEY1, and NOTCH1, as well as distal regions of DLL4 and EEG2. The top six enriched motifs in these DARs (KLF15, NRF1, ZBTB14, EGR1, SP4, and SP9) correspond to transcription factors involved in cell recruitment, gene regulation, cell differentiation and mitochondrial biogenesis. Similarly, DARs associated with NOTCH genes were identified in pDCs, although the differences were less pronounced (**Supplementary Figure 5A-B, Supplementary Table 5**). The top six enriched motifs in pDCs (NRF1, KLF15, KLF2, EGR1, ZBTB14, and KLF14) were also implicated in cell recruitment, gene regulation and cell differentiation (**Supplementary Figure 5C**). Since NOTCH signaling can mediate cell survival and proliferation in CD4+ TEM cells and memory B cells, we investigated whether enhanced chromatin accessibility could be observed in downstream targets within these cells, potentially explaining the enrichment of these gene expression pathways. Indeed, both CD4+ TEM cells and memory B cells exhibited higher peaks, indicating increased chromatin accessibility in HES1 (**Figure 4D+F**) and ETS2 (**Figure 4E+G**). The enhanced chromatin accessibility, primarily in promotor regions of NOTCH genes in DCs, aligns with increased transcription and may suggest prior training of these cells by the first dose, although this cannot be definitively confirmed.

**Figure 4.**
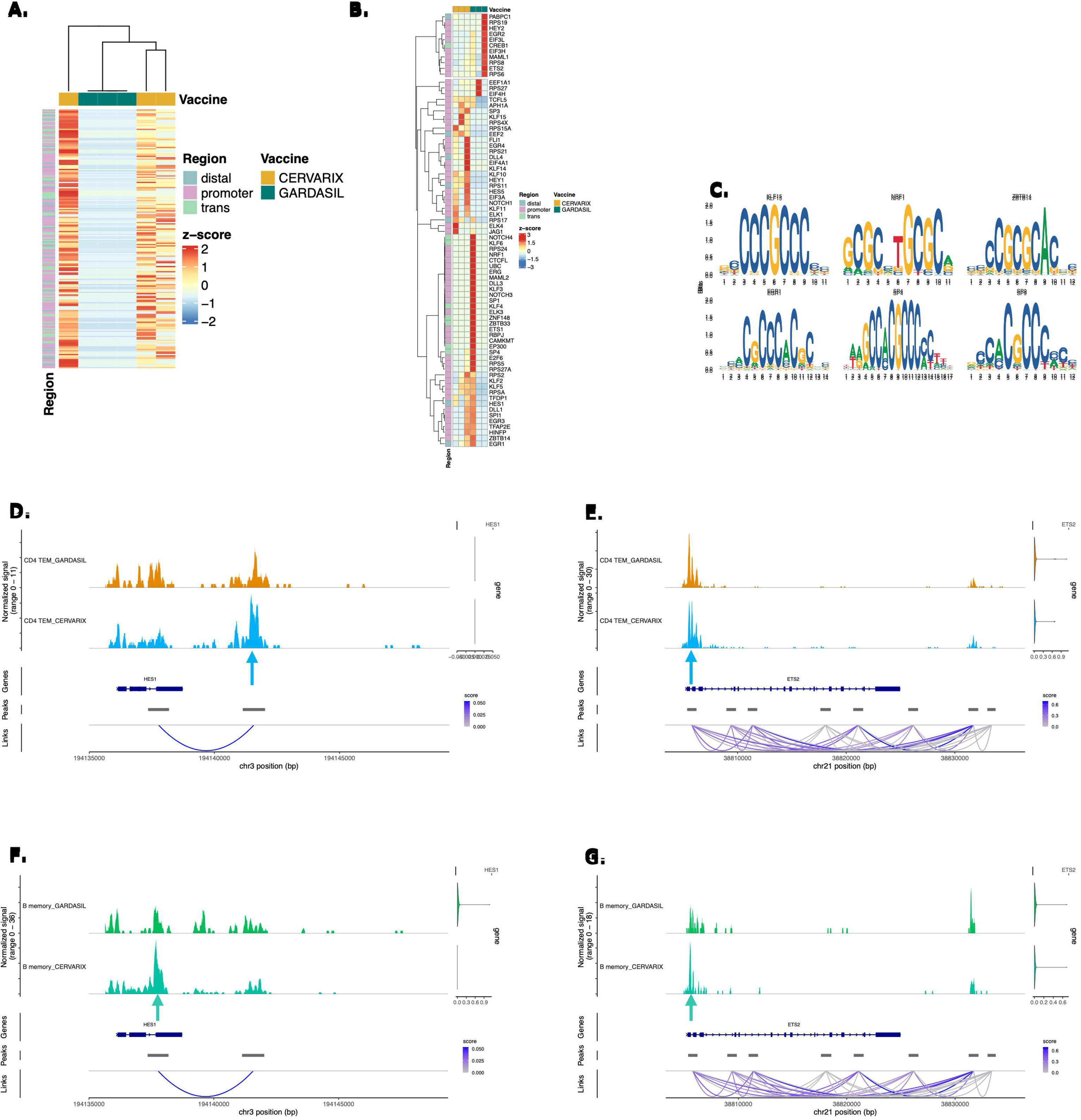
Cervarix enhances chromatin accessibility of NOTCH genes in cDC1. **A.** Heatmap of normalized accessibility at the top 200 DARs in cDC1 for each subject. Regions were classified as: promoter −2,000 bp to +500 bp; distal −10 kbp to +10 kbp – promoter; trans < −10 kbp or > +10 kbp. **B.** Heatmap of normalized accessibility of NOTCH-related DARs in cDC1 for each subject. **C.** The top enriched motifs identified from DARs in cDC1. **D-G.** Chromatin accessibility at the HES1 (**D+F**) or ETS2 (**E+G**) locus in CD4+ TEM (**D-E**) or memory B cells (**F-G**) grouped per vaccine. The coverage tracks represent the aggregate signal of transposase-accessible regions, with peaks indicating open chromatin regions. Gene expression levels are overlaid on the right. The plot extends 500 bp upstream and 10 kb downstream of the gene to capture potential regulatory elements. Links represent predicted chromatin interactions within the region.

### Th2 cells are activated and stimulate B memory cells and IgG4 production

Since the NOTCH signaling pathway, which is enriched in cDC1, appears to play an important role, we investigated whether this also leads to the activation of Th2 cells, as NOTCH signaling is a hallmark feature of these cells. To explore this, we first examined associations between cytokine concentrations and neutralizing antibody titers to identify potential skewing of CD4+ T cell responses. Overall, using Pearson correlations and clustering of pairwise distances, we observed that the neutralizing antibodies against HPV types tended to cluster into closely related types, with different cytokines showing either positive or negative correlations. Notably, IL4 was found to be positively associated with higher neutralizing antibody titers against HPV6, HPV16, HPV31, HPV32, and HPV58 (**Figure 5A**).

**Figure 5.**
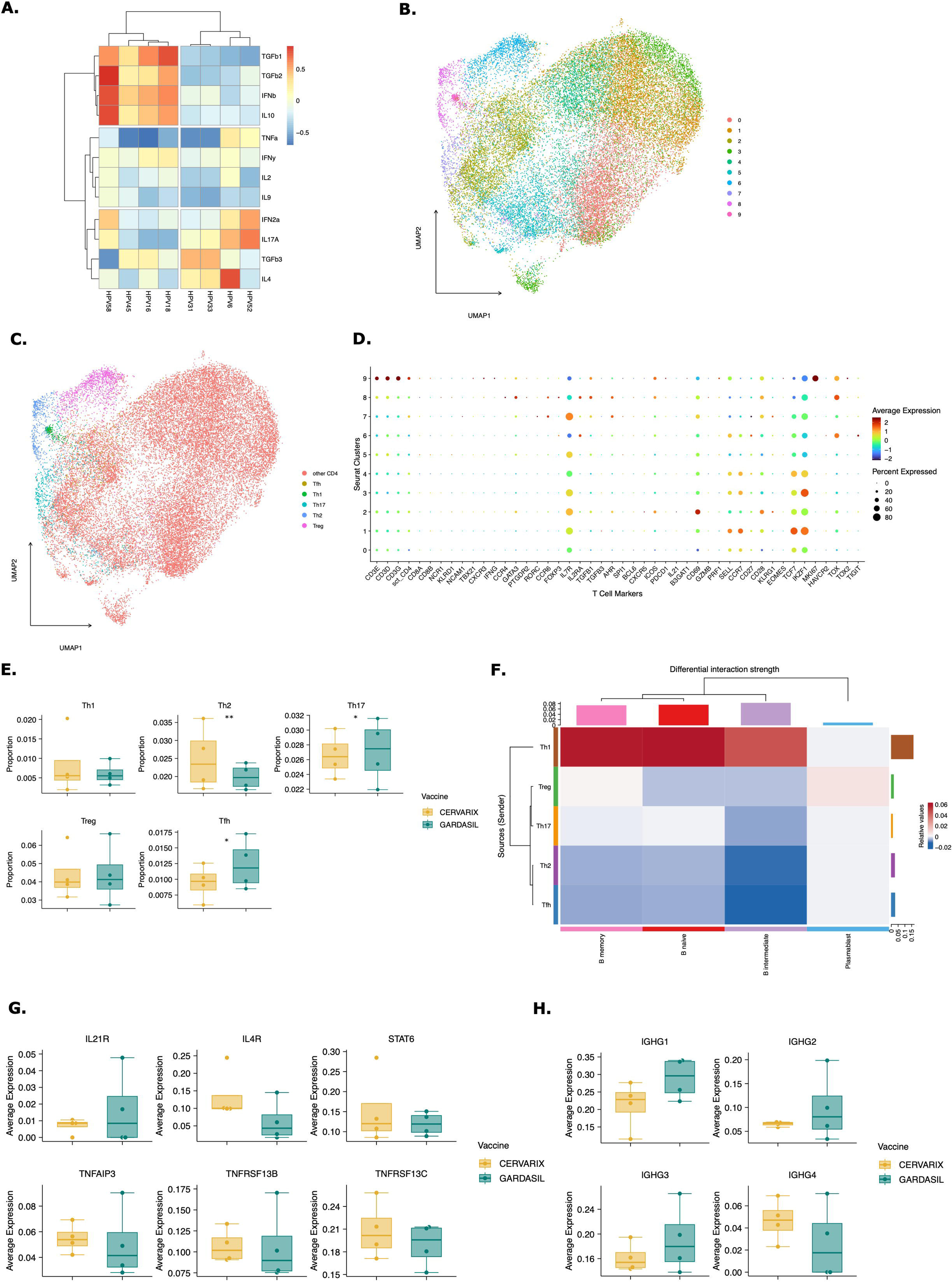
Enhanced Th2 cell help to B memory cells for improved affinity maturation. **A.** Correlation between cytokine concentrations and HPV-type specific neutralizing antibody titers. Color represents the Pearson correlation statistic. **B.** UMAP projecting displaying all CD4+ T cells after dimensionality reduction and clustering. Ten distinct clusters were identified. **C.** UMAP projection illustrating the manual annotation of T helper subsets. **D.** Dot plot showing the average expression of T cell markers across clusters. Based on marker expression, clusters were manually annotated. **E.** Boxplots representing the median (IQR) relative frequencies of CD4+ T cell subsets, stratified by vaccine (orange: Cervarix, green: Gardasil-4). **F.** Heatmap showing the differential strength of interactions between cell types. Rows represent sender cells (y-axis), and columns represent receiver cells (x-axis). Red indicates stronger signaling in Cervarix group, and blue indicates stronger signaling in Gardasil-4 group. The heatmap is clustered by the differential interaction strength, with bar plots on the rows and columns indicating the weight of signals sent or received, respectively. **G.** Boxplots showing the median (IQR) of expression of Th2 signaling genes in B memory cells, stratified by vaccine. H. Boxplots showing the median (IQR) of average expression of Immunoglobulin-related genes in B memory cells, stratified by vaccine. Abbreviations: Tfh: follicular T helper cell; Treg: regulatory T helper cell. * p-value <0.05, ** p- value <0.01, *** p-value <0.001 by Wilcoxon rank test.

To investigate whether Cervarix promotes greater Th2 differentiation, potentially facilitating extended affinity maturation and IgG4 production, we re-clustered all CD4+ T cells (**Figure 5B**). For each cluster, the average expression of common T cell markers was plotted (**Figure 5D**), and clusters were manually annotated as Th1, Th2, Th17, follicular helper T cells (Tfh), regulatory T cells (Treg), or other CD4+ T cells (**Figure 5B**).

A significant increase in the relative frequency of Th2 cells was observed on day 7 post-dose 2 of Cervarix compared to Gardasil-4 [OR: 1.42; 95%CI: 1.09-1.86], along with a significant decrease in Tfh and Th17 cells [OR: 0.61; 95%CI: 0.39-0.94] and [OR: 0.74; 95%CI: 0.56-0.97] respectively. No notable difference in Th1 cell frequencies was observed (**Figure 5E, Supplementary Table 6**).

Next, we examined whether Th2 cells provided enhanced help to B memory cells, as their relative frequency was also increased after Cervarix vaccination. Differences in communication between Th cells and B cells between the two vaccines were again assessed using the Cellchat package. This analysis revealed stronger interactions between Th2 cells and B memory, B intermediate and B naive cells after Cervarix compared to Gardasil-4 (**Figure 5F**). Conversely, Th1 responses were proportionally stronger after Gardasil-4, correlating with a classical anti-viral response (**Figure 5F**). Notably, T cell help after Gardasil-4 was more strongly directed towards plasmablasts (**Figure 5F**).

Interestingly, average IGHG4 gene expression, which drives IgG4 production, was elevated on day 187, compared to Gardasil-4, while other immunoglobulin types did not show similar increases (**Figure 5H**). This was accompanied by increases in average expression of genes related to Th2 signaling to B cells, such as STAT6, BAFFR, IL4R and IL21R, although it did not reach statistical significance after adjustment (**Figure 5G**).

### NOTCH signaling, cell cycling and survival gene signatures are associated with antibody breadth in other vaccines

Lastly, we aimed to investigate whether the gene signature identified here, including NOTCH signaling and increased cell survival, could be associated with breadth of the antibody response to other vaccines. The Gene Expression Omnibus (GEO) database was searched for publicly available gene expression data that also included antibody titers across multiple strains or types of the pathogen. Two relevant studies were identified (**Figure 6A**). The first study assessed a conjugated polysaccharide vaccine against meningococcus, with serum bactericidal antibody titers available for MenA and MenC (22). The second study examined the gene signatures of an mRNA COVID-19 vaccine and its relationship with neutralization antibody titers against two SARS-CoV-2 strains (23). For both studies, titers across strains were summed, and subjects were classified into high and low breadth groups based on the median value. Indeed, the top 20 significantly enriched pathways in high-breadth versus low-breadth responders after meningococcal vaccination included cell cycling and RNA translation processes (**Figure 6B, Supplementary Table 7**). After COVID-19 vaccination, the top 20 significantly enriched pathways included B cell receptor activation and cell migration pathways (**Figure 6C**). Furthermore, almost all the significantly enriched pathways after Cervarix identified in our dataset were also enriched in high-breadth responders for both vaccines (**Figure 6D**). These findings partially validate our results in this small population.

**Figure 6.**
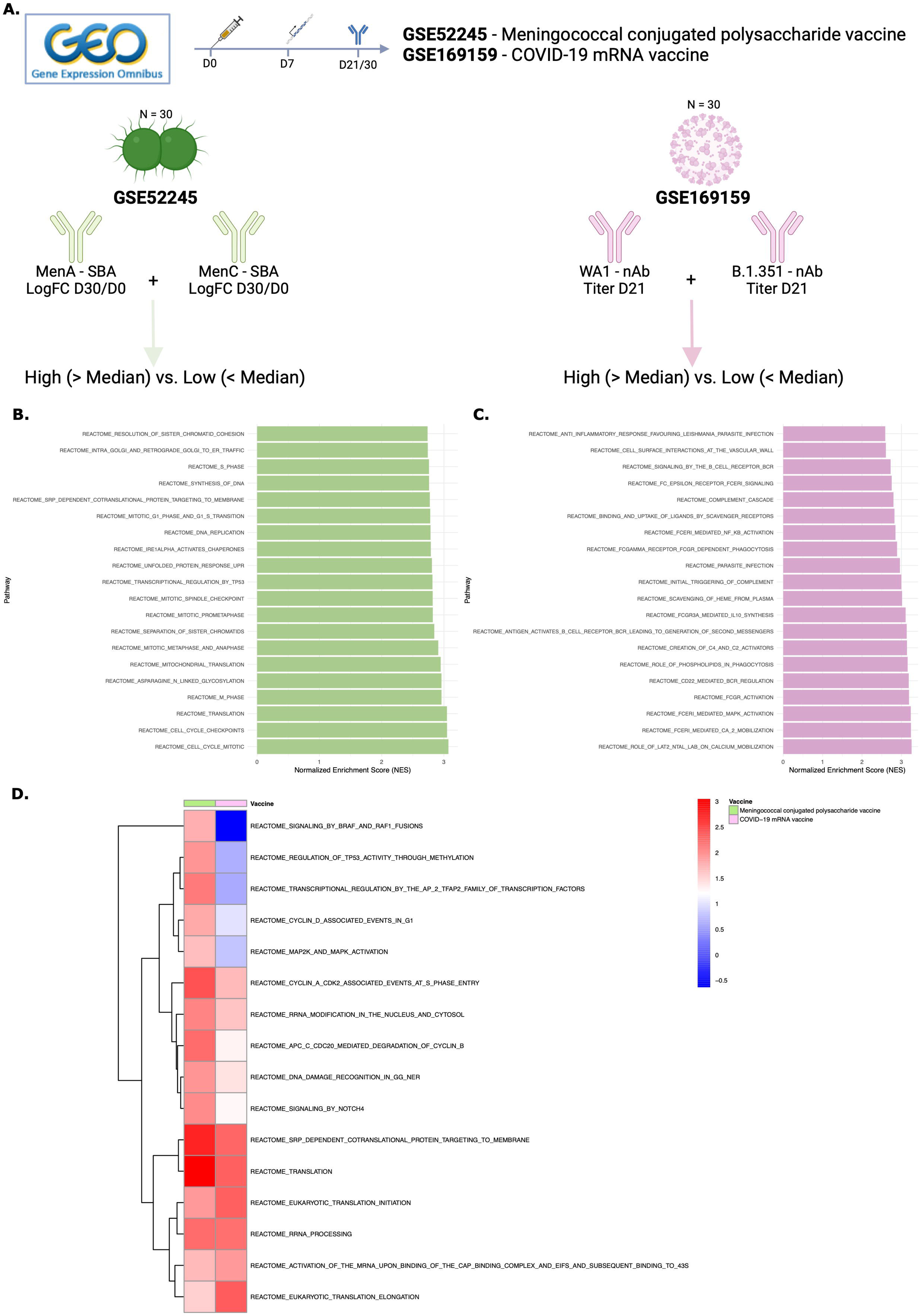
NOTCH signaling and cell cycling and survival gene signatures are associated with antibody breadth in other vaccines. **A.** The Gene Expression Omnibus (GEO) database was searched for publicly available gene expression data that included antibody titers across multiple strains or pathogen types. Two relevant studies were identified. The first examined a conjugated polysaccharide vaccine against meningococcus, with serum bactericidal antibody titers available for MenA and MenC. The second study analyzed gene signatures of an mRNA COVID-19 vaccine and their relationship with neutralization antibody titers against two SARS-CoV-2 strains. For both studies, titers across strains were summed, and subjects were categorized into high and low breadth groups based on the median value. **B-C.** Bar charts displaying the top 20 enriched pathways in high- vs. low-breadth subjects for GSE52245 (**B**) and GSE169159 (**C**). **D.** Heatmap showing the NES of pathways commonly enriched in both vaccines, clustered per vaccine. Abbreviations: NES: normalized enrichment score; GEO: gene expression omnibus; nAB: neutralizing antibodies; SBA: serum bactericidal assay.

## Discussion

In this study, we analyzed blood samples collected before first vaccination and 7 days after the the second vaccine dose administered to six homozygotic twin sisters. Our aim was to investigate the molecular mechanisms by which Cervarix, containing the adjuvant system AS04, induces a stronger cross-protective antibody response to oncogenic HPV types not included in the vaccine compared to Gardasil-4. Both vaccines generated high neutralizing antibody titers against HPV16 and HPV18. However, Cervarix elicited significantly higher neutralizing antibodies against HPV18 and HPV45, with a trend toward higher titers against HPV31, 33, 45, 52 and 58, compared to Gardasil-4.

At the cellular level, this response was associated with an increased relative frequency of DCs (pDC, cDC1, cDC2), and CD4+ TEM cells and memory B cells. Functionally, we observed enhanced DC recruitment and NOTCH signaling by cDC1, leading to active cell proliferation and survival of memory CD4+ T cells and B cells, even seven days after the second dose. These findings correlated with higher neutralizing antibody titers and a broader humoral response. This was accompanied by heightened cell-cell communication between Th2 cells and B memory cells, which stimulated the expression of cell survival genes, leading to increased IgG4 expression and potentially facilitating prolonged affinity maturation.

Consistent with previous research, our findings confirm that Cervarix induces higher neutralizing antibody titers against HPV18 and closely related HPV45 as compared to Gardasil-4 (15, 18). Studies attribute this superior response to the enhanced immunogenicity of the AS04 adjuvant (24, 25). AS04 has been shown to increase both the magnitude and durability of immune responses induced by HPV and hepatitis B vaccines (26). In preclinical models, AS04 was shown to activate APCs through TLR4, without directly targeting CD4+ T cells or B cells (20). Our study corroborates these findings by showing recruitment of cDC1 and pDCs which play a pivotal role in bridging innate and adaptive immunity. The activation of Toll-like receptor 4 (TLR4) by AS04 accounts for these robust DC responses, as TLR4 stimulation is well-recognized for enhancing antigen presentation and subsequent T-cell activation (27).

The upregulation of NOTCH-related genes in cDC1 cells in our dataset further suggests a role for NOTCH signaling in shaping adaptive immunity via antigen-presenting cells. Indeed, it has been shown that NOTCH signaling optimizes cDC1 differentiation in vitro, leading to better antigen presentation (28). In mice, NOTCH-ligand expressing DCs were found to better regulate T-cell effector functions leading to reduced tumor growth (29). Moreover, it was shown that NOTCH1 signaling increased antigen responsiveness in CAR-T cells (30). NOTCH signaling has also been shown to be critical for the differentiation of B cells to antibody-secreting cells, as its’ signaling enhances CD40 expression, as was seen in our cell-cell communication analyses, thereby supporting long-term humoral immunity (31). NOTCH signaling is well known to contribute to both T and B cell activation and differentiation (31–33). Thus, the contribution of NOTCH signaling to higher cell survival and proliferation on day 187, could allow extended time for affinity maturation, enhancing the vaccine’s capacity to generate cross-protective responses against related HPV types. This nuanced immune modulation by Cervarix may be a critical factor differentiating its overall immunogenicity profile.

Notably, gene regions associated with NOTCH signaling also exhibited greater accessibility in cDC1, indicating that the first dose of AS04 or prior antigenic encounters may have epigenetically primed these cells, which could contribute to enhanced responsiveness upon vaccination. Although, this aligns with other adjuvants such as AS03, inducing trained immunity (34), we cannot confirm this due to the lack of baseline samples. However, such epigenetic modifications may facilitate a more robust adaptive immune response, potentially explaining the differential vaccine-induced memory T and B cell activation observed in our study.

While the role of Th1 and pro-inflammatory responses in antiviral immunity is well-established, our findings demonstrate that AS04-driven Th2 activation and enhanced memory cell recruitment offer a complementary pathway for achieving cross-protection. This aligns with previous reports indicating a Th2-skewed immune response (35). The upregulation of Th2 cells is consistent with Th2-mediated responses observed with other adjuvants, such as MF59, and vaccines (26, 36, 37). Moreover, the robust interaction between Th2 cells and B cells, highlighted by our cell-cell communication analysis, underscores the critical role of Th2-dependent immune pathways. This process may enhance the breadth of neutralizing antibody responses and distinguish Cervarix from Gardasil-4.

Additionally, systems serology analyses have highlighted the importance of qualitative antibody attributes, such as Fc-mediated effector functions, in determining vaccine efficacy (19, 38, 39). Compared to Alum, AS04 has been shown to enhance antibody avidity, Fc-receptor functions, and memory B cell recall, although to a lesser extent than some other adjuvant systems (19, 38). More recently, studies have demonstrated that both Cervarix and Gardasil-4 elicit robust Fc-effector functions. However, Cervarix was found to coordinate these responses more effectively, resulting in higher antibody-dependent complement activating responses (39).

Cervarix’s ability to promote a Th2-biased response and sustained cell proliferation leads to an increased IgG4 production. IgG4 antibodies are produced following prolonged antigen recognition and, although they have limited Fc-receptor functionality, they possess important immunomodulatory effects (40). While the induction of IgG4 has been associated with weaker vaccine-specific immunity in the context of COVID-19 vaccinations (41), this does not appear to be the case for HPV vaccines.

Lastly, the robust immune response against HPV18 observed for both vaccines may have clinical implications, given the association of HPV18 with aggressive cervical cancer phenotypes. Higher titers of cross-neutralizing antibodies, particularly against HPV18-related types, are linked to reduced rates of persistent infection and cervical intraepithelial neoplasia (42).

This study leveraged single-cell RNA sequencing to provide a detailed view of cell-type-specific immune responses. The analysis of transcriptional changes offered insights into the molecular pathways activated by Cervarix. The use of homozygotic twins minimized genetic variability, enabling precise comparisons and enhancing the reliability of observed differences. However, some limitations remain. The twin-based design, while strengthening internal validity, involved a small sample size (n = 12), limiting generalizability of the findings. However, the validation in public gene expression data confirmed the correlation of these pathways with breadth of antibody responses. Moreover, several studies found an association of gene signatures related to cell cycling and durable antibody responses (34, 43, 44), including the tremendous effort of defining a transcriptional atlas of 13 vaccines (45, 46). Immune responses were assessed only seven days after the second vaccination, and baseline cellular immune responses were not measured. While all subjects were HPV naive with no expected pre-existing immunity, a baseline PBMC sample would have allowed identification of trained immunity and of vaccine-specific changes over time. Future longitudinal studies are needed to evaluate antibody persistence and long-term immune memory. Moreover, newer HPV vaccines, such as Gardasil-9, may elicit different and broader immune responses, warranting comparative analysis. Other limitations of this study are that neutralizing antibody titers were the sole functional immunological parameter assessed, and that cellular immune responses have not been confirmed using conventional flow cytometry. As a result, the findings reflect overall changes in the PBMC compartment and cannot be directly extrapolated to vaccine- or antigen-specific cellular immune responses.

In conclusion, AS04 enhances DC recruitment, with cDC1 in particular exhibiting increased NOTCH signaling, evidenced by elevated gene expression and epigenetic modifications likely induced by prior training. This signaling promoted cell survival, RNA translation and proliferation, particularly in memory B cells up to seven days post-vaccination. Additionally, AS04 broadened T cell help, including enhanced Th2 responses. The resulting adaptive immune response relied more on memory T and B cells with higher cell survival. This ‘paced’ immune activation supports more effective maturation of adaptive immune cells, potentially explaining the enhanced cross-reactivity observed with Cervarix. These findings underscore the critical role of adjuvant selection in optimizing vaccine efficacy and highlight the potential of AS04 to drive robust, long-lasting immunity.

---

**Supplementary Figure 1.**
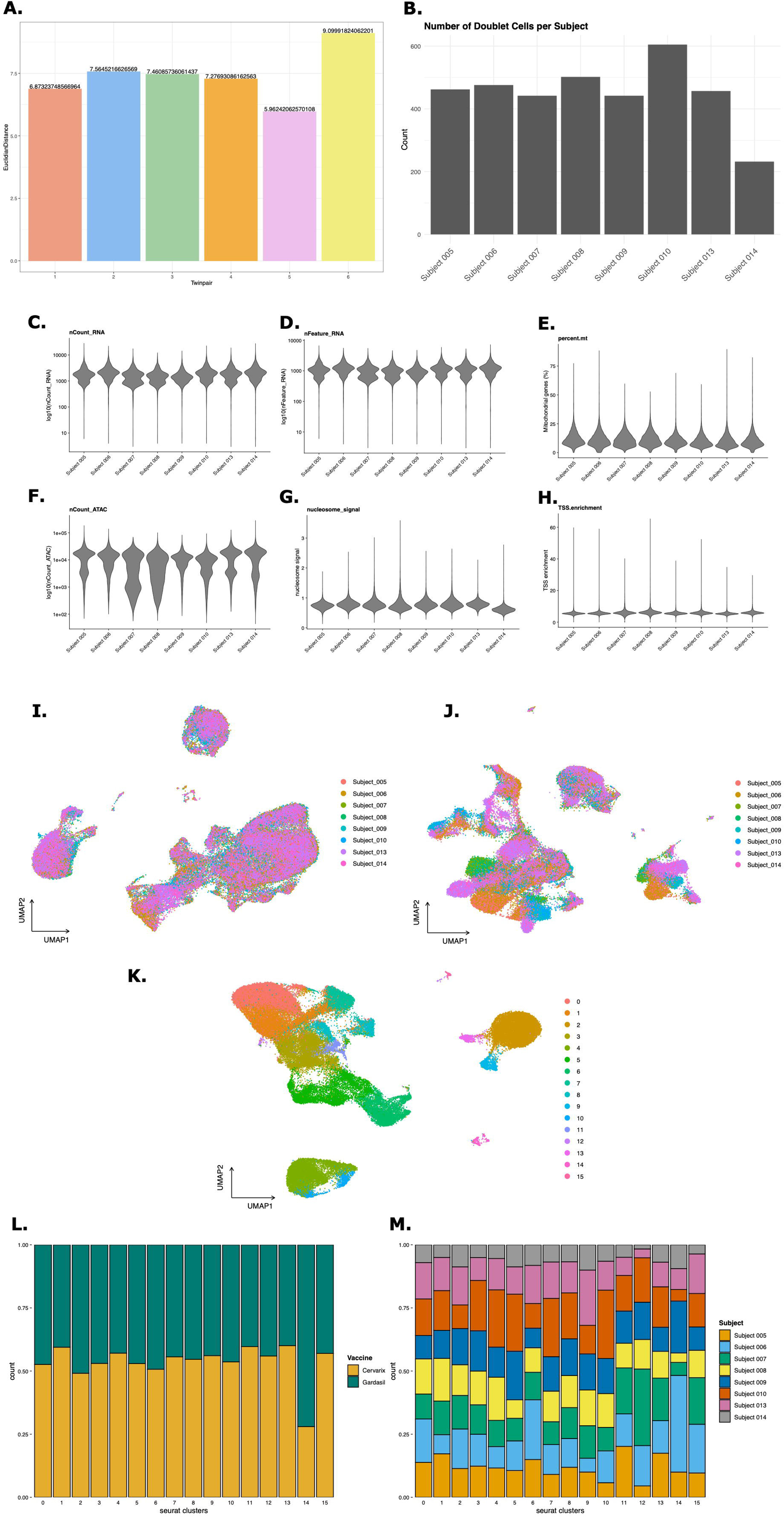
Single cell RNA and ATAC sequencing quality. **A.** Euclidian Distances between twin sisters based on neutralizing antibody titers for all HPV types. **B.** Bargraph showing the number of doublet cells for each subject identified using the DoubletFinder package. **C-H.** Violin plots showing the distribution of six quality metrics across all cells. Quality metrics were number of reads (**C**), number of transcripts (**D**), percentage of mitochondrial genes (**E**), nmver of ATAC reads (**F**), nucleosome signal (**G**), and TSS enrichments score (**H**). **I.** UMAP projection of gene expression data. **J.** UMAP projection of ATAC data. **K.** Combined UMAP projection. Both gene expression and ATAC data have been integrated using Harmony integration to remove batch effect. A combined UMAP projection has been made using weighted neighbor clustering. **L.** Per-cluster cell proportions from both vaccines. **M.** Per-cluster cell proportions from each subject.

**Supplementary Figure 2.**
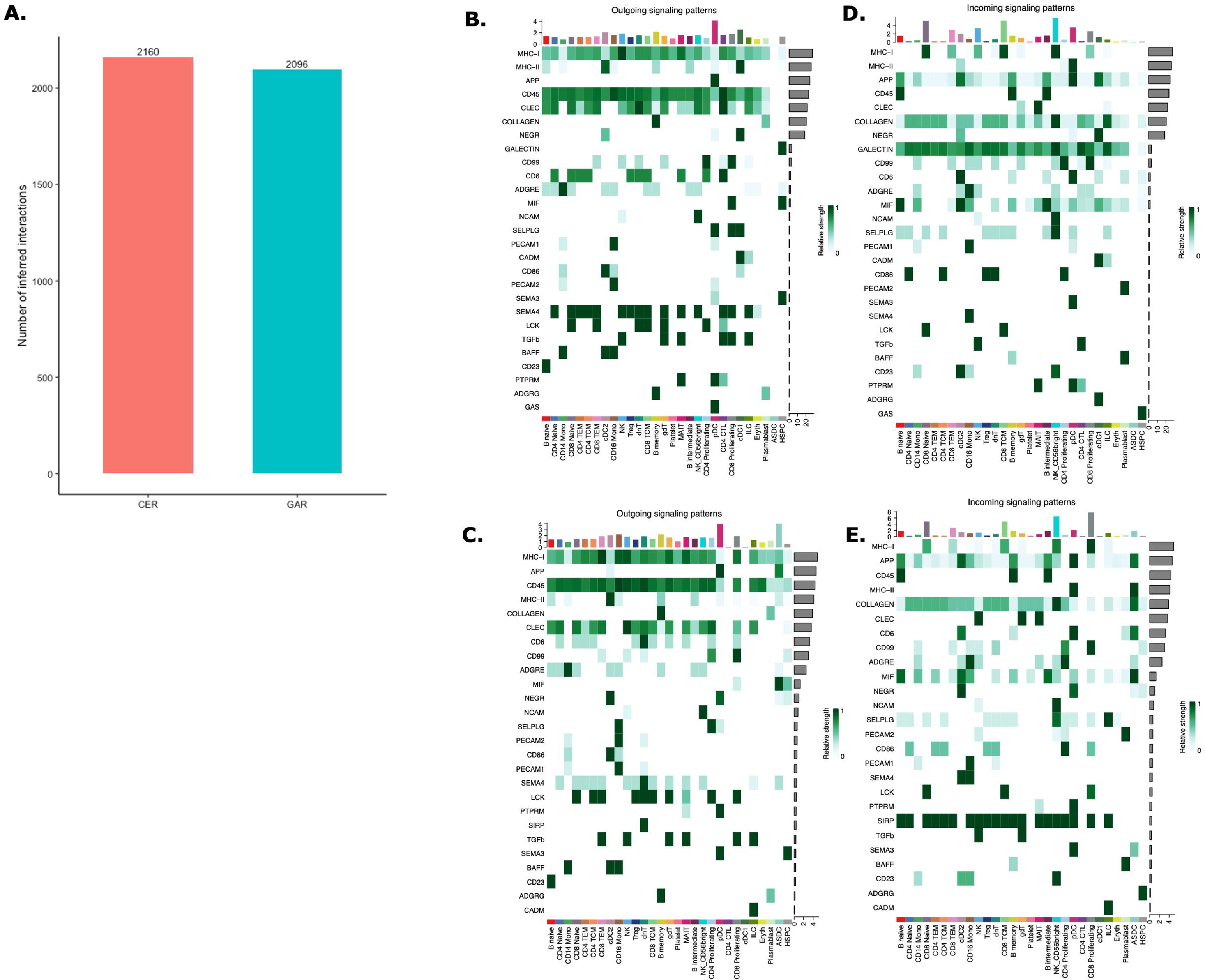
Cell-Cell communication analysis. **A.** Bar plot showing the total number of interactions per vaccine. **B-C.** Heatmaps showing outgoing signals for each cell type in Cervarix (**B**) and Gardasil-4 (**C**). “Outgoing” refers to signals sent by the specified cell types. Signals are ranked by ‘importance’ and the extent to which cells utilize them. For Cervarix, MHC-I and MHC-II are the two most prominent outgoing signals, predominantly sent by DCs, whereas MHC-II is comparatively less important in Gardasil-4. Bar plots on the columns represent the total number of signals sent by each cell type, while the bar plots on the rows indicate the total number of signals sent overall. Green highlights the relative strength of the outgoing signal. **D-E.** Heatmap showing incoming signals for each cell type in Cervarix (**D**) and Gardasil-4 (**E**). G-J. NicheNet analysis showing signaling from DCs to CD4+ TEM and memory B cells. **F.** Ligand activity plot highlighting the most important ligands sent by cDC1 and pDC to CD4+ TEM cells. **G.** Target gene heatmap. Expression of genes in CD4+ TEM cells (columns) that are influenced by the top ligands (rows). **H.** Ligand activity plot highlighting the most important ligands sent by cDC1 and pDC to memory B cells. **I.** Target gene heatmap. Expression of genes in memory B cells (columns) that are influenced by the top ligands (rows). Abbreviations: CD14 Mono: classical CD14+ monocytes; CD16 Mono: non-classical CD16+ monocytes; NK: Natural Killer cells; ASDC: AXL+ Siglec-6+ dendritic cells; cDC1/cDC2: conventional dendritic cells type 1 or type 2; pDC: plasmacytoid dendritic cell; CD4 TEM: CD4+ T effector memory cells; CD4 TCM: CD4+ T central memory cells; Treg: regulatory T cells. CD4 CTL: CD4+ cytotoxic T cell; CD8 TEM: CD8+ T effector memory cells; CD8 TCM: CD8+ T central memory cells; ILC: innate lymphoid cells; gdT: gamma delta T cells; dnT: double negative T cells; AUPR: area under the precision recall curve; LFC: log fold change; CER: Cervarix; GAR: Gardasil-4.

**Supplementary Figure 3.**
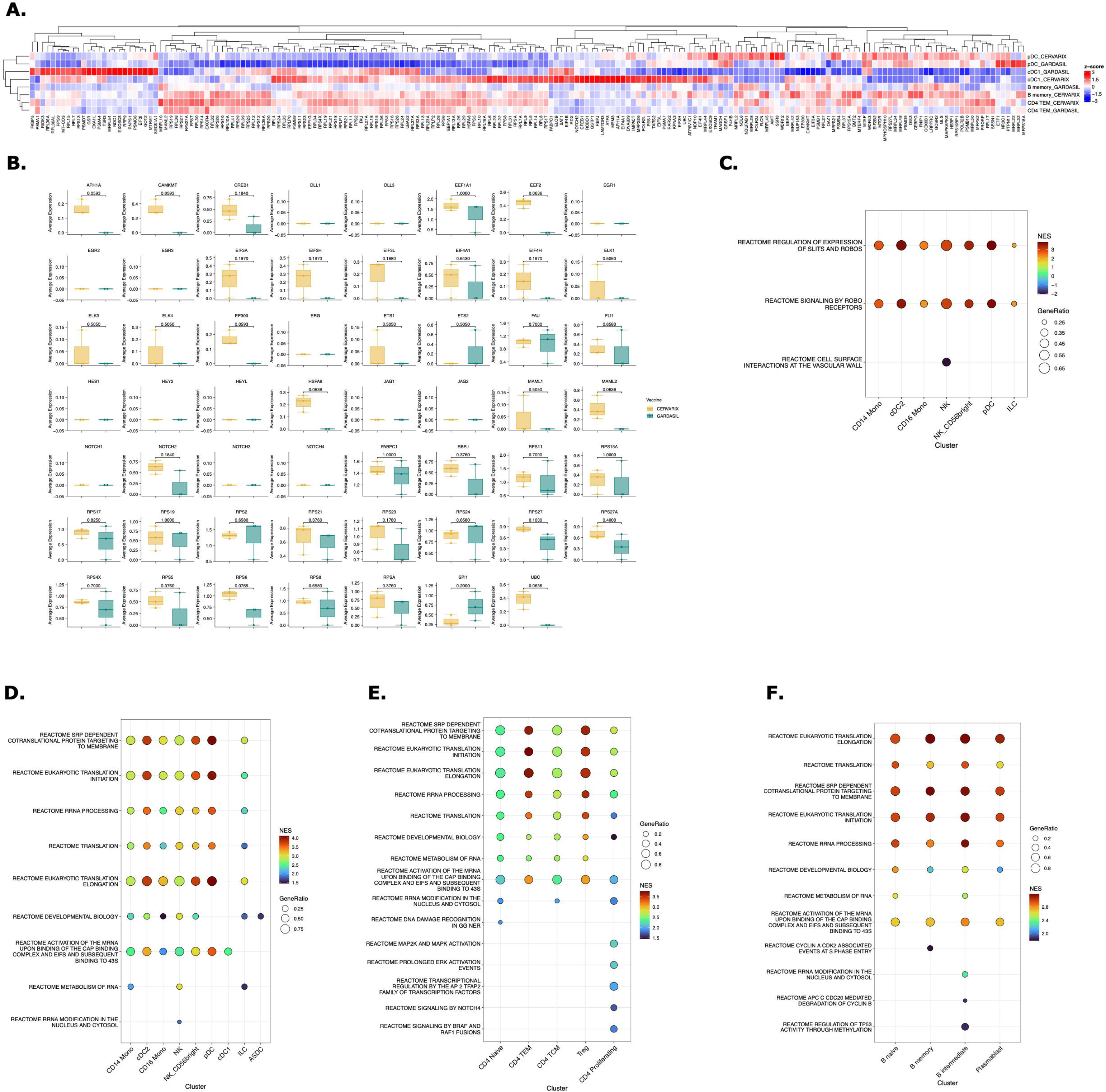
Gene set enrichment analysis in all cells. **A.** Clustered heatmap of all genes involved in identified enriched pathways in cDC1, pDC, CD4+ TEM, and B memory cells. Color represents column-wise z-scores. **B.** Boxplot showing the average gene expression of NOTCH-related genes in cDC1, grouped by vaccine. **C.** Dot plot illustrating pathways related to cell migration in innate immune cells, highlighting upregulated pathways (yellow and red) and downregulated pathways after Cervarix compared to Gardasil-4. **D-F.** Dot plot illustrating pathways related to RNA translation, cell metabolism, and proliferation in innate immune cells (**D**), CD4+ T cells (**E**) and B cells (**F**), with upregulated pathways after Cervarix compared to Gardasil-4. Abbreviations: CD14 Mono: classical CD14+ monocytes; CD16 Mono: non-classical CD16+ monocytes; NK: Natural Killer cells; ASDC: AXL+ Siglec-6+ dendritic cells; cDC1/cDC2: conventional dendritic cells type 1 or type 2; pDC: plasmacytoid dendritic cell; CD4 TEM: CD4+ T effector memory cells; CD4 TCM: CD4+ T central memory cells; Treg: regulatory T cells. CD4 CTL: CD4+ cytotoxic T cell; CD8 TEM: CD8+ T effector memory cells; CD8 TCM: CD8+ T central memory cells; ILC: innate lymphoid cells; gdT: gamma delta T cells; dnT: double negative T cells; NES: normalized enrichment score. * p-value <0.05, ** p-value <0.01, *** p-value <0.001 by Wilcoxon rank test.

**Supplementary Figure 4.**
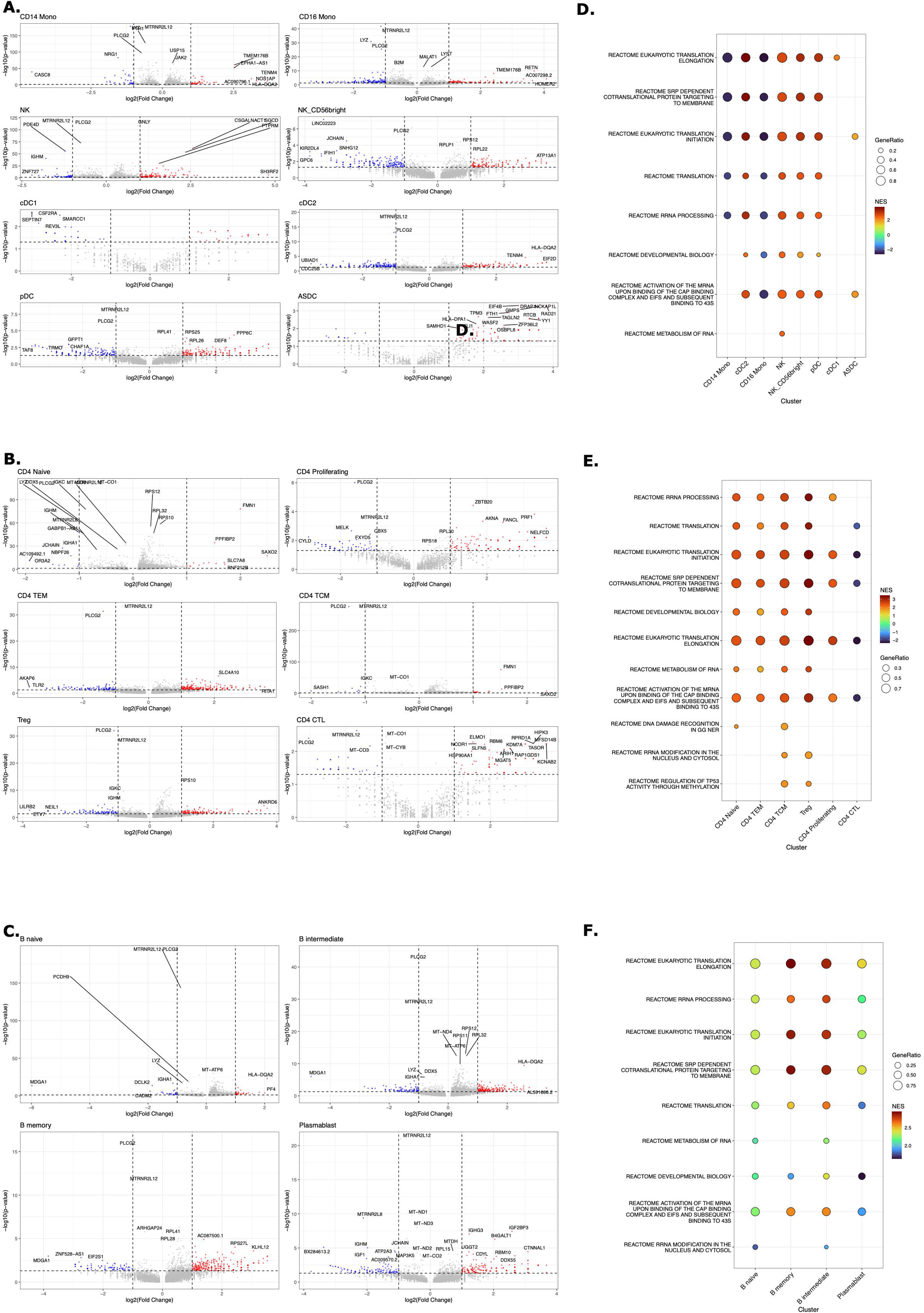
Gene expression correlation with breadth of neutralizing antibody response. A-C. Volcano plots of differentially expressed genes (DEGs) for innate cells (**A**) CD4+ T cells (**B**) and B cells (**C**) of subjects with high breadth of neutralizing antibody titers (higher than median of the sum of titers of all types) compared to cells of subjects with low breadth of neutralizing antibody titers (lower than median of the sum of titers of all types). **D-F.** Enriched pathways related to cell cycling and RNA translation in innate immune cells (**D**), CD4+ T (**E**), and B cells (**F**). Data are stratified by subjects with a high breadth of neutralizing antibody titers (greater than the median sum of titers across all HPV types) versus those with a low breadth of neutralizing antibody titers (less than the median sum of titers across all HPV types). Abbreviations: CD4+ T cells and B cells after Cervarix compared to Gardasil-4. CD14 Mono: classical CD14+ monocytes; CD16 Mono: non-classical CD16+ monocytes; NK: Natural Killer cells; ASDC: AXL+ Siglec-6+ dendritic cells; cDC1/cDC2: conventional dendritic cells type 1 or type 2; pDC: plasmacytoid dendritic cell; CD4 TEM: CD4+ T effector memory cells; CD4 TCM: CD4+ T central memory cells; Treg: regulatory T cells. CD4 CTL: CD4+ cytotoxic T cell; CD8 TEM: CD8+ T effector memory cells; CD8 TCM: CD8+ T central memory cells; ILC: innate lymphoid cells; gdT: gamma delta T cells; dnT: double negative T cells; NES: normalized enrichment score.

**Supplementary Figure 5.**
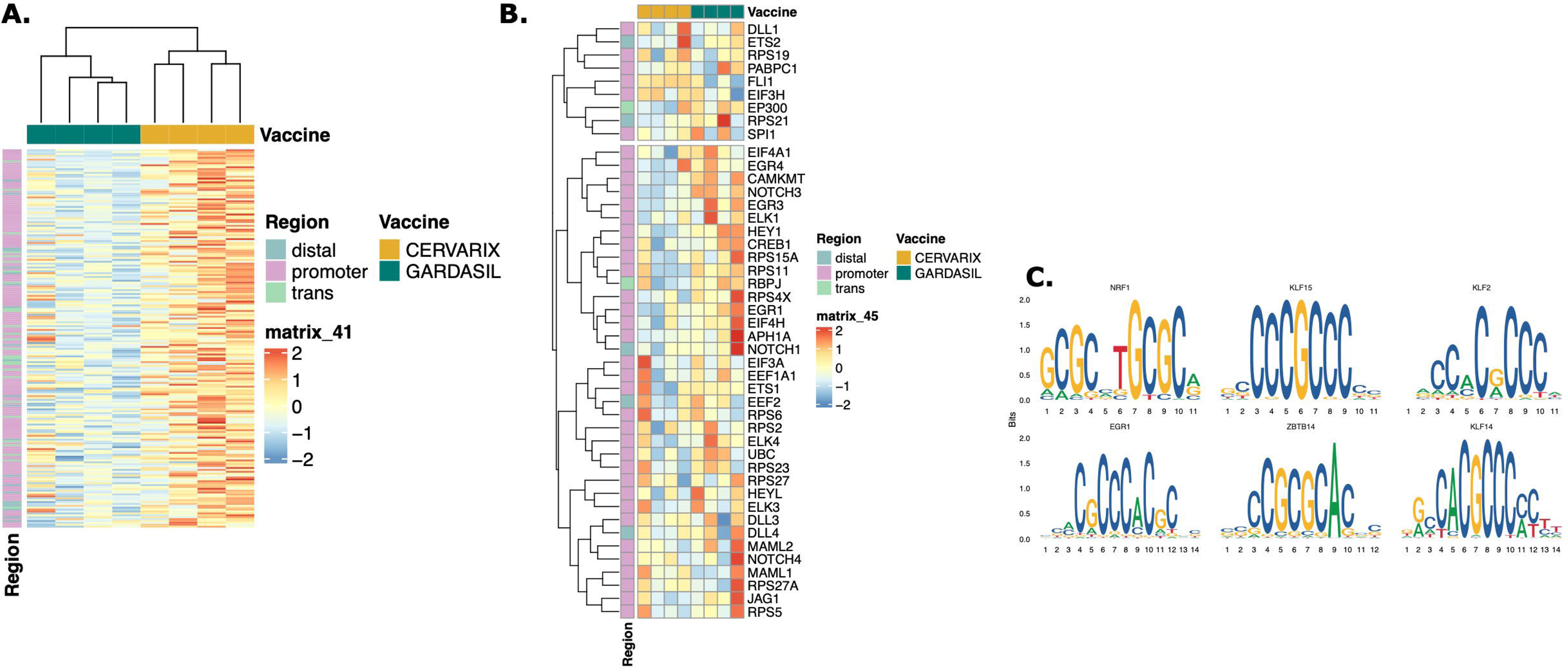
Epigenetic remodeling and chromatin accessibility in pDC. **A.** Heatmap showing normalized chromatin accessibility at the top 200 DARs in pDCs for each subject. Regions were classified as follows: promoter −2,000 bp to +500 bp; distal −10 kbp to +10 kbp – promoter; trans < −10 kbp or > +10 kbp. **B.** Heatmap displaying normalized accessibility of NOTCH-related DARs in pDCs for each subject. **C.** Top enriched motifs identified from DARs in pDCs.

## Methods

### Study Design

A blinded randomized interventional study was performed at the Center for Vaccinology (CEVAC, Ghent University and Ghent University Hospital) in Ghent, Belgium between June 2014 and April 2015. Participants were homozygous female twins between 9 and 13 years that were eligible for HPV vaccination according to national recommendations. All girls were healthy, virgin and did not have prior exposure to HPV or any HPV vaccine or vaccine containing AS04. The primary objective of the study was to investigate molecular mechanisms of cross-neutralizing properties of Cervarix® compared to Gardasil-4®. Gardasil-4 was used, as Gardasil-9 had not yet been introduced at the time of the study. All study procedures adhered to ICH and GCP guidelines. The study was approved by the Ethics Committee of Ghent University Hospital and by the Belgian Federal Agency for Medicines and Health Products (FAHMP)(EudraCT: 2013-002340-90, NCT 01914367). Informed consent was obtained from all participants and from both parents.

### Study Vaccines

Each sister per twin pair was randomized to receive either Cervarix or Gardasil-4. Both vaccines are recombinant vaccines consisting of virus-like particles (VLP) containing L1 proteins of HPV. Cervarix targets HPV types 16 and 18 and uses Adjuvant System 04 (AS04, Al(OH)3 + TLR4 agonist monophosphoryl Lipid A (MPL)). Gardasil-4 targets HPV types 6, 11, 16 and 18 and is formulated with amorphous AlHO9PS-3 adjuvant. For both vaccines, two doses were administered intramuscularly to all participants, 6 months apart.

### Sample collection

Ten mL of blood were collected by venous puncture in serum separation blood collection tubes (Becton Dickinson Vacutainer tubes) from all participants at baseline (pre-dose 1, day 0) and 7 days after the second dose (day 187). Serum was seperated after centrifugation for 10 minutes at 1300-2000g and stored in 500 μL aliquots at -20°C for the determination of neutralizing antibody titers and cytokines. At day 187, 60 mL of blood was collected in lithium-heparin coated tubes (Becton Dickinson Vacutainer tubes) for the isolation of peripheral blood mononuclear cells (PBMCs). After 1:2 dilution in Hanks buffered salt solution (HBSS), PBMCs were isolated by density gradient centrifugation (Lymphoprep™), washed twice in HBSS, suspended in freezing solution (10% dimethyl sulfoxide/90% fetal bovine serum v/v), frozen at a concentration of up to 10 million cells/mL and stored in liquid nitrogen.

### Pseudovirion-based neutralization assay

Neutralizing antibodies against HPV types 6, 16, 18, 31, 33, 45, 52 and 58 were determined at day 0 and day 187 by pseudovirion-based neutralization assay (PBNA) as described previously (21, 47–49). Briefly, pseudovirions were produced in HEK293TT cells and purified by ultracentrifugation in an Optiprep gradient. Pseudovirions comprise HPV L1 and L2 proteins that encapsidate a Gaussia luciferase reporter plasmid. Expression of Gaussia luciferase is quantified by luminescent reaction with the luciferase substrate coelenterazine after transduction of the plasmid into HeLaT reporter cells by pseudovirion infection. The pseudovirion infection is blocked and the the transduction of reporter genes is reduced in the presence of neutralizing antibodies. Serum samples were serially diluted in 3.33-fold increments to achieve a final dilution of 1:40 to 1:180 000 on the neutralization assay. Antibody titers were calculated as serum dilutions inhibiting 50% of the luciferase activity (EC50 value). EC50 values greater than 40 were defined as neutralizing antibody-positive. Sufficiently active HPV11 pseudovirions were not available for this study.

### Cytokine assay

The Meso Scale Multi-Array Technology (Meso Scale Discovery) was used for measurement of cytokine levels. A cytokine panel containing the following analytes was screened: IL2, IL4, IL9, IL10, IL17A, TNFa, IFN2a, IFNb, IFNy, TGFb1, TGFb2, TGFb3, using 25lμl of each serum from each donor in duplicates. Samples were randomized to avoid batch effects. Results were extrapolated from the standard curve from each specific analyte and plotted in picograms per milliliter, using the DISCOVERY WORKBENCH v.4.0 software (Meso Scale Discovery).

### Single cell RNA and ATAC sequencing

Single cell multiome (RNA+ATAC) sequencing (Chromium Next GEM Single Cell Multiome ATAC + Gene Expression, Document Number GC000338 Rev F, 10X Genomics) was done using PBMCs collected at day 7. Nuclei were prepared from thawed PBMCs according to 10X Genomics demonstrated protocol (Nuclei Isolation for Single Cell Multiome ATAC + Gene Expression Sequencing, Document Number CG000365 Rev C, 10x Genomics). Frozen cells were thawed and incubated with DNase. Thawed cells were counted, and the viability was determined by staining the cells with Trypan blue. Cell suspensions were lysed to obtain isolated nuclei, according to the manufacturer’s instructions. Briefly, cells are incubated with lysis buffer on ice for 4 minutes, in alignment with previous optimization. Nuclei were washed, resuspended, and the percentage of dead cells was determined by incubating the nuclei with trypan blue and counting using an automated cell counter. Nuclei morphology was determined by staining the nuclei with Hoechst, and nuclei are classified as A – D (A, smooth, uniformly round nuclei with well-resolved edges; B, mostly intact nuclei with minor evidence of blebbing; C, nuclei with ruffled edges; D, nuclei no longer intact.). Only samples with the majority of nuclei type A and absence of nuclei type D are used in the subsequent steps of the protocol. Single Cell Multiome ATAC and Gene Expression (GEX) libraries were prepared using the Chromium Single Cell Multiome ATAC + Gene Expression platform (10X Genomics, Pleasanton, CA). 10,000 nuclei were targeted for each sample. Isolated nuclei were transposed and partitioned into Gel Beads-in-emulsion (GEMs) using the 10x Chromium Controller and Next GEM Chip J. GEMs were visually inspected, and only samples with an opaque and uniform aspect were used for library preparation. ATAC and GEX libraries were generated from the same pool of pre-amplified transposed DNA/cDNA. Representative traces and quantitation of both libraries were determined using Bioanalyzer High Sensitivity DNA Analysis (Agilent, Santa Clara, CA). Sequencing was done on Illumina NovaSeq S4 targeting 20,000 reads per nucleus for gene expression and 25,000 reads per nucleus ATAC-seq.

Fastq files were processed 10x Genomics Cell Ranger v5.0.1 using 10x Genomics Cloud Analysis (50, 51). Reads were mapped to the GRCh38 human reference genome and counted without depth normalization. The filtered count matrix was then analyzed using the Seurat (v5.1.0)(52) and Signac packages (v1.14.0)(53) in R. Low quality cells were identified and removed based on the following criteria: more than 100 RNA unique molecular identifiers, less than 25% mitochondrial read fraction, transcription start site (TSS) enrichment score of more than 2 and more than 200 ATAC fragments in peaks. Doublet cells were identified and removed using the DoubletFinder package in R (54). Peak calling was performed using the CallPeaks function. Data normalization, dimensional reductio and batch correction using Harmony integration was done independently on RNA and ATAC assays, which were then integrated using weighted nearest neighbors method (WNN). Cells were clustered using the Louvain algorithm using the integrated Uniform Manifold Approximation and Projection (UMAP), and global differences between clusters were assessed using Principal component analysis (PCA).Cell annotation was carried out using the reference expression dataset derived from Azimuth (55, 56). Differential Gene Expression per cell type between the two vaccine conditions was analyzed using the FindMarkers function (test.use = ‘wilcox’) in the Seurat package and visualized on volcano plots. The obtained sets of DEGs were also used for hierarchical k-means clustering and z-scaled averaged gene expression per subject for each cell type separately were visualized in heatmaps using pheatmap. Ranked DEGs per cell type were subjected to Gene Set Enrichment analysis (GSEA) using the clusterProfiler package in R (57). Reactome pathways were used (58). Average gene expression of significant pathways was calculated using AddModulescore and were correlated to neutralizing antibody titers. Cell-Cell communication analyses were done using the R packages CellChat (59) and NicheNet (60). A per-cell motif binding site activity score is calculated using chromVAR, utilizing a collection of 746 transcription factors from the JASPAR database. To assess chromatin accessibility changes between the two vaccine conditions, differentially accessible regions (DARs) were identified using the FindMarkers function with the LR test for ATAC data in Signac. Regions with significant differences in accessibility were visualized using heatmaps to highlight vaccine-specific regulatory elements. Motif enrichment analysis was performed on DARs using FindMotifs, identifying overrepresented transcription factor binding motifs that may play a role in vaccine-induced immune responses. The top enriched motifs were visualized using MotifPlot, displaying sequence logos that represent nucleotide conservation at these regulatory elements. To further explore transcriptional regulation, CoveragePlot was used to integrate chromatin accessibility and gene expression at key loci. ATAC-seq peaks at this locus suggested potential regulatory elements and links between regulatory regions and gene promoters were overlaid to infer chromatin interactions.

### Statistical analysis

Geometric means of neutralizing antibody titers were calculated for each vaccine at each timepoint. Log10 transformed titers are presented in boxplots (median and IQR). The Mann-Whitney U test was used to evaluate differences in neutralizing antibody titers at day 187. To assess the overall difference in neutralizing antibody titers of all HPV types between twin sisters, the Euclidian distance was calculated as follows:

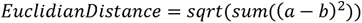

Where a and b are the neutralzing antibody titers for each HPV type.

Cytokine concentrations were log10-transformed and tested for significant differences using the Kruskal-Walis test. Pearson correlation coefficients were calculated to evaluate associations between cytokine levels and HPV-type specific neutralizing antibody titers. Differences between the vaccines in relative frequencies of different cell types were analyzed using the Wilcoxon rank-sum test with Bonferoni correction for multiple comparisons. Differentially expressed genes were identified using the Wilcoxon rank-sum test and p-values were adjusted for multiple testing using the Benjamini-Hochberg false-discvery rate (FDR) procedure. Statistical significance was defined as an adjusted p-value ≤ 0.05 and a fold-change > 1. Correlation analyses between module scores of genes in enriched pathways and HPV neutralizing antibody titers were conducted using Kendall’s rank correlation coefficients using the cor.test function in R. Hierarchical clustering was performed on the correlation matrix to group similar rows based on pairwise distances. The Euclidean distance was computed using the dist function, and the hclust function with the complete linkage method was applied for clustering. Continuous data are presented as mean (±SD) or median (IQR), while categorical data are shown as N (%). P-values < 0.05 were considered statistically significant. All analyses were done using R and R Studio.

## Acknowledgements

VDO has received a travel grant (V476023N) from FWO (Flanders Research Foundation) for his stay at Emory University to conduct the single-cell RNA sequencing experiments and analyses. We thank all the participants in the study, the clinical and laboratory staff of CEVAC.

## Author contributions

V.DO.: data curation, funding acquisition, formal analysis, investigation, methodology, project administration, resources, visualisation, writing, review, and editing. AC. S: data curation, investigation, methodology, project administration, validation, review, and editing. M.P.: data curation, investigation, methodology, project administration, validation, review, and editing. G.W.: investigation, project administration, review, and editing. A.W.: investigation, writing, review, and editing. F.DB.: project administration, review, and editing. P.S.: data curation, investigation, methodology, project administration, validation, review, and editing. T.W.: data curation, investigation, methodology, project administration, validation, review, and editing. I.L.-R.: conceptualization, funding acquisition, investigation, methodology, supervision, review, and editing. A.S. data curation, formal analysis, investigation, methodology, supervision, review, and editing. RP.S.: conceptualization, data curation, formal analysis, funding acquisition, investigation, methodology, project administration, supervision, validation, review, and editing. G.L.-R.: conceptualization, data curation, formal analysis, investigation, methodology, supervision, validation, review, and editing. All authors have read and agreed to the published version of the manuscript.

## Conflicts of Interest

G.L.-R. provided consulting services and received consulting fees from Virometix, Osivax, ICON Genetics, Sumitomo Pharma, Curevo, and Minervax. I.L.-R. declares that her institution received funding from GSK, Icosavax, Virometix, Janssen Vaccines, Curevac, Moderna, Osivax, MSD, ICON Genetics, and OSE Immunotherapeutics for other vaccine trials; and from Janssen Vaccines and MSD for consulting services. All others declare no conflicts of interest.

## Data Availability

The data that support the findings of this study are available on request from the corresponding author [VDO]. The data are not publicly available due to them containing information that could compromise research participant privacy.

## Code Availability

The R code to generate the results presented here will be available on Github (centerforvaccinology) after peer review.

## Notes

### Clinical Trial

NCT01914367

### Author Declarations

The study was approved by the Ethics Committee of Ghent University Hospital and by the Belgian Federal Agency for Medicines and Health Products (FAHMP)(EudraCT: 2013-002340-90, NCT 01914367).

